# Delayed Healthcare Seeking and Associated Factors for Common Childhood Illnesses among Caregivers with Under-Five Children in Southwestern Ethiopia, 2023

**DOI:** 10.1101/2024.01.12.24301231

**Authors:** Gamechu Atomsa Hunde, Kalkidan Fikadu, Tigist Demeke

## Abstract

**Background:** For under-five children, receiving timely and appropriate medical attention is crucial in preventing serious and fatal complications. Unfortunately, evidence shows that parents of young children frequently delay seeking care, contributing to the death of many kids before they even get to a medical facility.

**Objectives:** The study aimed to assess delay in healthcare seeking and associated factors for common childhood illnesses among caregivers with under-five children visiting Yem special woreda public health facilities, 2023.

**Methods:** A facility-based cross-sectional study was conducted among 333 caregivers of under-five children diagnosed with common childhood illnesses visiting Yem special woreda public health facilities. Systematic random sampling was employed and data collection was carried out using an interviewer-administered questionnaire. Delay was characterized as a long time (typically >24 hours) between disease onset and start of the necessary treatment. Data was entered using Epi Data version 4.7 and exported to Statistical Package for the Social Sciences version 25.0. Bi-variable and multivariable logistic regression analyses were conducted to identify the factors that influence the delay in healthcare seeking. Adjusted odds ratios with a 95% confidence interval were used to determine the associations. Statistically significant variables were identified based on a p-value < 0.05.

**Results:** A total of 326 caregivers participated in the study with a response rate of 98%. The proportion of delayed health care seeking was 74.5%. Child ≥ 12 months (AOR =1.99, 95% CI: 1.11-3.57), rural residence (AOR = 2.41, 95% CI: 1.35-4.28), no community health insurance (AOR = 1.91, 95% CI: 1.07-3.42), traditional treatment (AOR = 2.98, 95% CI: 1.46- 6.10), and self-medication at home first (AOR = 2.73, 95% CI: 1.32-5.63) and perceiving illness as mild (AOR= 2.64, 95% CI: 1.28-5.42) were factors associated with delayed healthcare seeking.

**Conclusion and recommendation:** The study showed delay in health care seeking for common childhood illnesses among caregivers was high. Hence, reducing delays necessitates the implementation of public education campaigns, collaboration with local organizations, and the provision of counseling for caregivers regarding childhood illnesses.

## INTRODUCTION

Childhood illnesses encompass any sickness, disability, or abnormality that occurs during the developmental period from fetus to adolescence (<u>1</u>). Common childhood illnesses are infectious diseases that are significant causes of morbidity and mortality in under five children worldwide, especially in low- and middle-income nations (<u>2</u>).

Malaria, pneumonia, and diarrhea are among the most common infectious diseases that exhibit a high susceptibility in young children and a prominent contributors to childhood mortality (<u>3</u>, <u>4</u>). Prevention of these diseases requires access to essential interventions like nutrition, vaccination, breastfeeding, hygiene, reducing air pollution, and affordable drugs. Additionally, effective treatments for pneumonia, malaria, and diarrhea include Amoxicillin tablets, artemisinin-based combination therapy (ACT), oral rehydration salts (ORS), and zinc respectively (<u>5-7</u>).

The burden of illness and subsequent death from treatable diseases can be reduced by timely medical care (<u>8</u>). Caregiver delays in seeking medical care can have detrimental effects on children’s health and diminish the safety and efficacy of medical interventions (<u>9</u>, <u>10</u>). This delay is often defined as a significant period, typically exceeding 24 hours, between the onset of illness and the commencement of essential treatment (<u>11</u>).

In order to enhance the accessibility of comprehensive primary healthcare and mitigate prevalent childhood illnesses, the World Health Organization (WHO) and the United Nations Children’s Fund (UNICEF) have collaboratively formulated the Integrated Management of Childhood Illness (IMCI) strategy (<u>12</u>). Ethiopia is currently incorporating these strategies within its primary healthcare and health extension programs (<u>13</u>).

Healthcare services for common childhood illnesses were usually underutilized and accessed with delays (<u>14</u>). The delay in accessing healthcare services encompasses challenges related to recognizing the disease, making decisions about seeking care and arranging transportation to healthcare facilities (<u>15</u>). Delayed healthcare for children’s illnesses, even when services are accessible, results in preventable illnesses, mortality, and economic burdens (<u>16</u>, <u>17</u>). The postponement of seeking medical care for children under five years old may lead to severe and potentially fatal complications (<u>18</u>, <u>19</u>). A significant number of under-five deaths are related to preventable or treatable conditions, and delayed healthcare seeking by caregivers can exacerbate these issues. A considerable proportion of childhood mortality is attributable to delays in accessing healthcare, indicating insufficient advancement in reducing child fatality rates (<u>20</u>).

In 2021, global under-five mortality reached 5.0 million, with significant disparities observed based on the child’s country of birth (<u>21</u>, <u>22</u>). Sub-Saharan Africa has the worst rate, registering 74 deaths per 1,000 live births, 15 times higher than Europe/North America and 19 times higher than Australia/New Zealand respectively (<u>21</u>).

Common childhood illnesses such as pneumonia, diarrhea, and malaria collectively accounted for nearly 30% of global under-five deaths in 2019, with Africa being responsible for a million deaths annually due to these conditions (<u>3</u>, <u>23</u>). In 2023, in Ethiopia, approximately 22.5% of children under five suffer from common childhood illnesses such as acute respiratory infections (ARI), diarrhea, and fever (<u>24</u>). Efforts to address this issue include improving healthcare accessibility, enhancing healthcare education for caregivers, and promoting early intervention and preventive care for children. Organizations and governments around the world continue to work towards reducing under-five mortality rates by addressing these and other related challenges (<u>25</u>).

While Ethiopia has made progress in reducing child mortality through various interventions, including immunization and disease prevention programs, high child mortality rates persist, especially among underserved populations (<u>26</u>, <u>27</u>). Ethiopia has set a target to reduce the child mortality rate to below 20 per 1,000 live births by the year 2035 (<u>26</u>).

Studies conducted across different nations indicate that mothers and primary caregivers tend to delay seeking healthcare for their children’s illnesses. In Cameroon, it was recorded that 88.1% of caregivers experienced a delay in seeking healthcare for common childhood illnesses (<u>28</u>). In Nepal, this delay was reported as 62.7% (<u>29</u>), while in Bhubaneswar, the delay was found to be 15.77% (<u>30</u>).

In Ethiopia, evidences shows a significant proportion of delay for common childhood illnesses, with proportions ranging from 86.3% in Jeldu district, Oromia region, 73.5% in Addis Ababa, and 73% in Northwest Ethiopia’s, Aneded district (<u>31-33</u>). Factors such as lack of access to healthcare, insufficient awareness of when to seek medical help, and economic barriers can all contribute to delays in seeking healthcare for children. Additionally, in some regions, cultural or social factors may also play a role in delaying healthcare seeking (<u>28</u>, <u>34</u>, <u>35</u>).

While studies have examined the delay in seeking care for common childhood illnesses in certain regions of the country (<u>34</u>, <u>35</u>), it is essential to understand the variations of each factor within different cultures, environments, and communities to enhance early healthcare practices (<u>36</u>). Thus, this study aimed to assess delays in healthcare-seeking and associated factors for common childhood illnesses among caregivers of under-five children in Yem special woreda public health facilities.

## METHODS AND MATERIALS

### Study area and period

The study was conducted from June 1-30, 2023 in Yem Special Woreda, situated approximately 297 kilometers southwest of Addis Ababa. It comprises one town administration, three municipalities, and 34 rural kebeles. Based on unpublished data obtained from the Yem special woreda finance and development office in 2022, the total population of the area is 116,044 individuals, including 18,244 children under the age of five.

The Woreda has one primary hospital (Saja Primary Hospital) which serves a population of 32,234 and five health centers/ HC (Fofa, Semonama, Toba, Deri, and Gesi) (<u>37</u>). The hospital operates with a total staff of 240 and offers a range of services, including one under-five outpatient unit (OPD), adult OPD, psychiatry OPD, emergency OPD, x-ray, ophthalmology OPD, ANC and family planning, delivery ward, neonatal intensive care unit (NICU), laboratory, ART and TB, pharmacy as well as adult and pediatric inpatient unit services. The HCs in the Woreda provide primary healthcare services in under-five OPD, adult OPD, Maternity and delivery, laboratory, and pharmacy units.

### Study Subjects and Procedures

The study employed a health facility-based cross-sectional study among caregivers of under-five children diagnosed with common childhood illnesses visited under-five OPD at Yem special woreda public health facilities during the study period. Caregivers who visit the health facilities with appointment or comeback for follow up without new complaint and whose child needed urgent referral was excluded from the study. The largest sample size for the study was estimated using single proportion formula assuming a 95% confidence level, margin of error of 5%, and a proportion p=73% from a study of delay in seeking healthcare behavior for common childhood illnesses in Aneded district, Northwest Ethiopia (<u>38</u>). Considering 10% non-response rate, the final sample size was 333. A systematic random sampling method was applied to select the eligible caregivers after proportionally allocating number caregivers to be selected from each health facility. Accordingly, 149 caregivers from Saja Hospital, 43 from Fofa health center, 37 from Semonama health center, 48 from Toba and 56 care givers from Deri health center were selected by systematic random sampling. The caregivers were interviewed keeping the interval of 2 between consecutive caregivers whose child was diagnosed with one of common childhood illness (pneumonia, malaria or diarrhea) by doctors/health care providers.

### Operational definitions

**Health care-seeking:** getting care from a qualified health care professional at government health centers or private hospitals/ clinics (39).

**Delay in healthcare-seeking:** care that is sought from medical facilities more than 24 hours after becoming aware of symptoms (31, 34, 35).

**Common childhood illnesses:** ARI, diarrheal disorders, and febrile illnesses are the common childhood illnesses in this study (31).

**First of choice treatment:** defined as the setting where a caregiver makes their initial contact after the child becomes unwell (health institutions, traditional healers, self-medication, religious places) (40).

**Self-medication:** is when mothers become aware of their children’s sickness, begin all available treatments, and apply them without a healthcare professional’s prescription (39).

**Traditional medicine:** traditional healers, wogesha, herbalists, and magicians (holy water) handle patients with apparent illnesses and sickness using experience-based knowledge and practice (39).

**Perceived severity of illness:** subjective assessment of the child’s level of discomfort by the caregiver; classified as mild, moderate, or severe based on the caregivers’ (29).

**Good Knowledge of general danger signs:** if mothers were able to mention more than three of the nine danger signs identified by the WHO such as not able to breastfeed/ unable to drink or eat, vomits everything, had convulsions/ convulsing now, lethargic or unconscious, fast breathing, severe chest in drawing, fever, low body temperature, movement only when stimulated or no movement even when stimulated otherwise it is poor (41, 42).

**Monthly income:** classified according to the Ethiopian civil service proclamation into three categories: low income (<3000 ETB), medium income (3000-7500 ETB), and high income (>7500 ETB) (43).

### Data collection Tool and procedures

The questionnaire was developed by reviewing relevant literature (<u>19</u>, <u>28</u>, <u>33</u>, <u>34</u>). A structured interview administered questionnaire was employed to collect the data. This questionnaire has 6 components: socio-demographic characteristics, caregiver’s knowledge and child’s illness-related factors, health care access-related factors, caregivers’ previous experience-related factors, decision makers’ behavior-related factors, and promptness of care seeking for illness. The questionnaire was prepared in English and translated to local languages Amharic and Yemissa. Face-to-face exit interview was conducted in a separated room by one Nurse who has Bachelor of Science holder in each health facility and supervised by two master’s holder nurses.

### Data processing, analysis, and presentation

The data was checked for completeness, consistency, and errors before proceeding to data processing and analysis. Data was entered into Epidata version 4.7 and then exported to SPSS software version 25.0 to conduct data cleaning and analysis. Descriptive analysis was performed to determine proportions, standard deviations, and frequencies. A binary logistic regression was used, and bi-variable and multivariable analysis was done. Variables with p-values less than 0.25 in the bi-variable analysis were taken to multivariable analysis. Model fitness was checked using the Hosmer–Lemeshow goodness of the model with p-value found to be greater than 0.05. An odds ratio with a 95% confidence interval with P-value <0.05 was used to determine the association between the independent and the outcome variables. Variables that are statistically significant at p-value < 0.05 were identified as factors for delay in health care seeking. Finally, the result of the analyses was presented in texts, tables, and charts accordingly.

### Data quality control

The questionnaire was translated into the local languages of Amharic and Yemissa by language experts and reviewed by subject matter experts. Then, the Amharic and Yemissa versions were translated back to English, to verify the consistency. Content validity was checked by the subject matter experts. A pretest was carried out with 5% of the sample size (17 caregivers) in Gesi HC a week before the actual data collection time and was excluded from the actual study. A one-day orientation was provided for data collectors and supervisors on the purpose of the study, how to approach study subjects, and how to use the questionnaire. The collected data were reviewed and checked for completeness everyday by the supervisors and principal investigator.

### Ethical consideration

Ethical approval was obtained from Institutional Review Board (IRB) of Jimma University Institute of Health with the letter Ref. No: JUIH/IRB/393/23. A formal letter for permission and support was obtained from Jimma University School of Nursing to the stakeholders of Yem special woreda health office. Letter of permission and support was written to each health facility from Yem special woreda health office. Verbal informed consent was taken from the study participants before starting the interview after were informed of the purpose, and benefits of the study. The respondents were notified that they have full right to refuse or terminate the questionnaire if they feel uncomfortable with the questions at any point and information from any respondent was kept anonymous and confidential. The collected data were stored in anonymous way in a secret and secure place. The data would not be used for another purpose apart from research purposes. Participation is completely voluntary basis and confidentiality of the information is guaranteed by a secret code and kept anonymously. The hard copy of the collected data was kept locked cabinet and the soft copy is secured by a password on the computer.

## RESULTS

### Socio-demographic characteristics of participants and children

In this study, out of 333 caregivers offered to the interview, 326 caregivers who had children under the age of five with common childhood illnesses completed the interview making a response rate of 98%. Among the participants, 186 (57.1%) were between the 25-34 age group, with a mean (SD) age of 31.03 (± 6.535) years. One hundred ninety-two (58.9%) of the children brought to the health facilities were 12 months and older, with a mean (SD) age of 19 (±13.704) months. The majority 309 (94.8) of the participants were married. More than half 171(52.5%) of the participants had attended primary school, and 155 (47.5%) identified as housewives by their occupation. In terms of income, 190 (58.3%) participants reported low monthly incomes (<3000 ETB). ***(Table 1)***

**Table 1:**
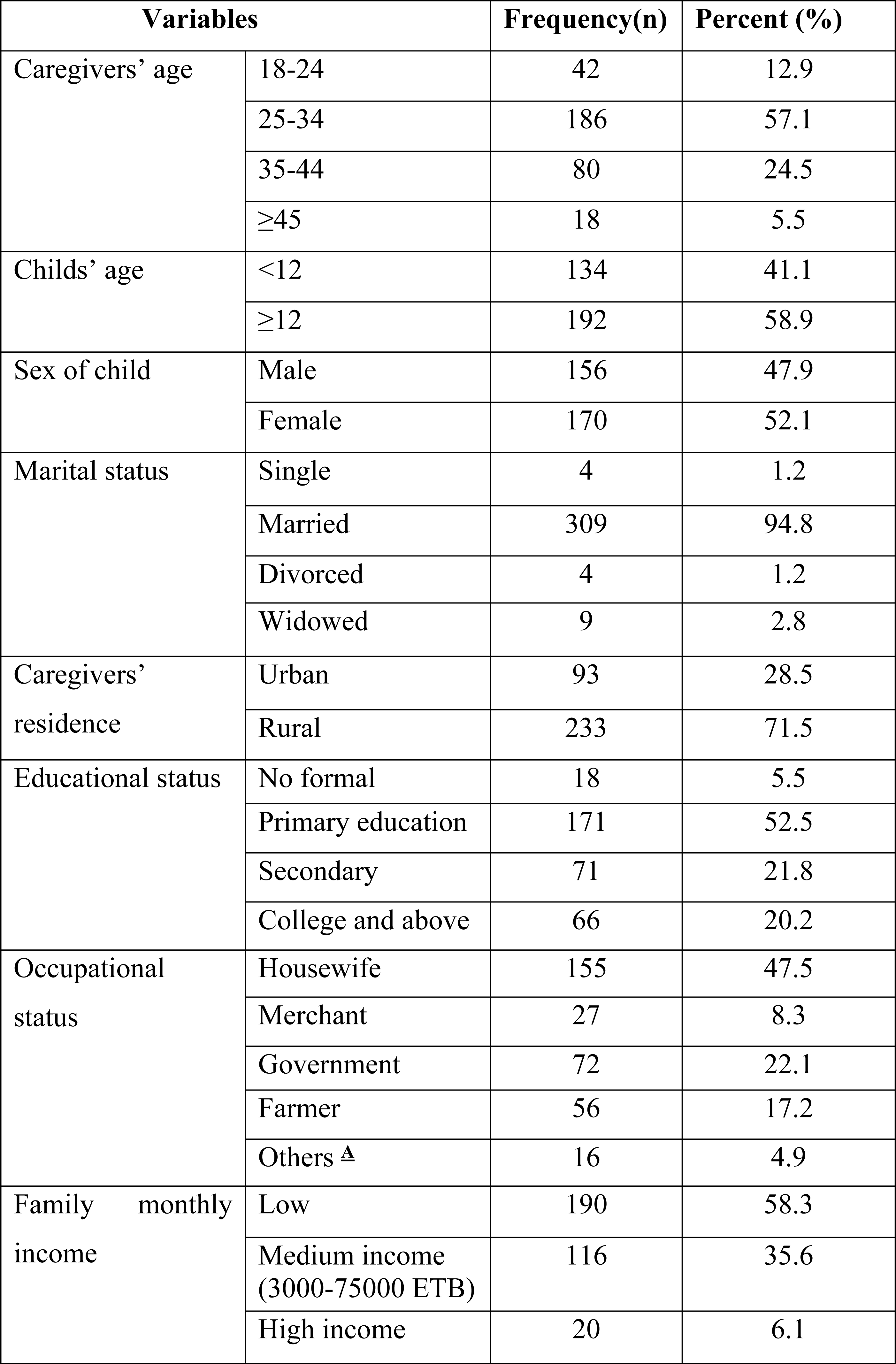
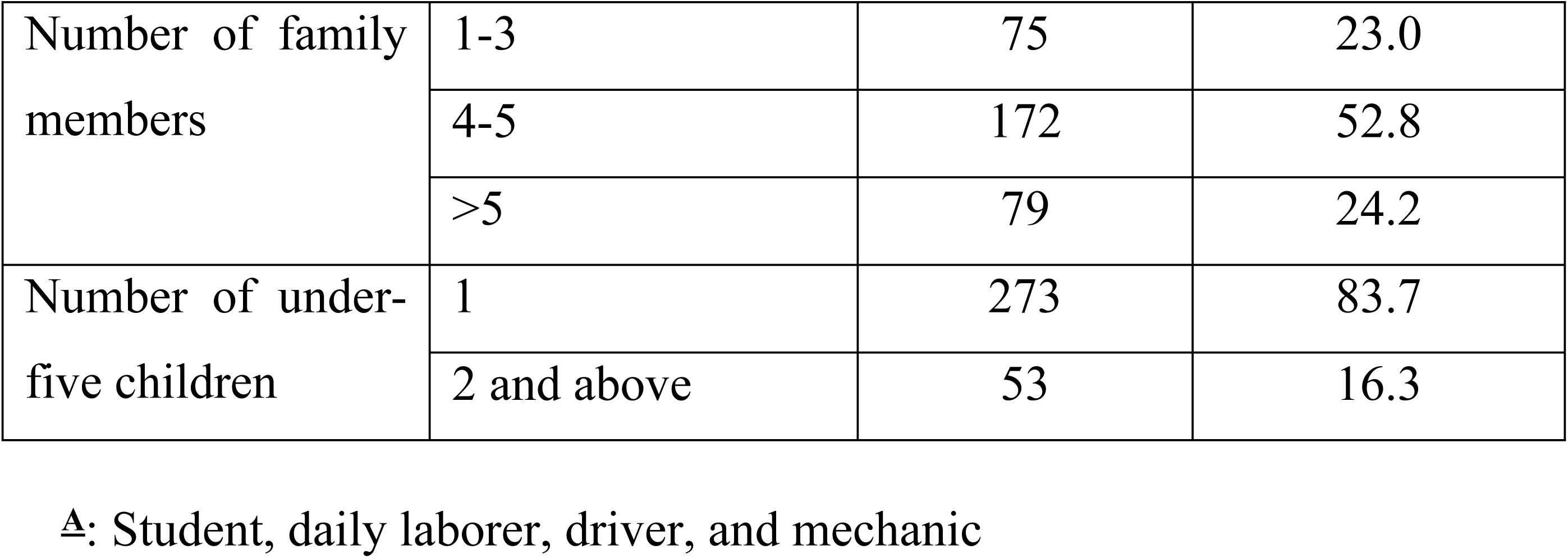
Socio-demographic characteristics of the study participants and their children in Yem special woreda public health facilities, Southwestern Ethiopia, 2023 (N=326).

### Caregivers knowledge and child-illness-related Factors

Among the total participants, the majority 278 (85.3%) of the participants had poor knowledge of the danger signs of common childhood illnesses. Regarding the children’s presenting symptoms, 183 (56.3%) had present with cough. Nearly one-third 104 (31.9%) of the participants perceived their child’s illness was caused by microorganisms. One hundred twenty-five (38.3%) of the study participants expressed a preference to seek initial medical care for their sick children at a health institution. More than one-third of participants 118 (36.2%) rated their child’s illness as moderate. ***(Table 2)***

**Table 2:**
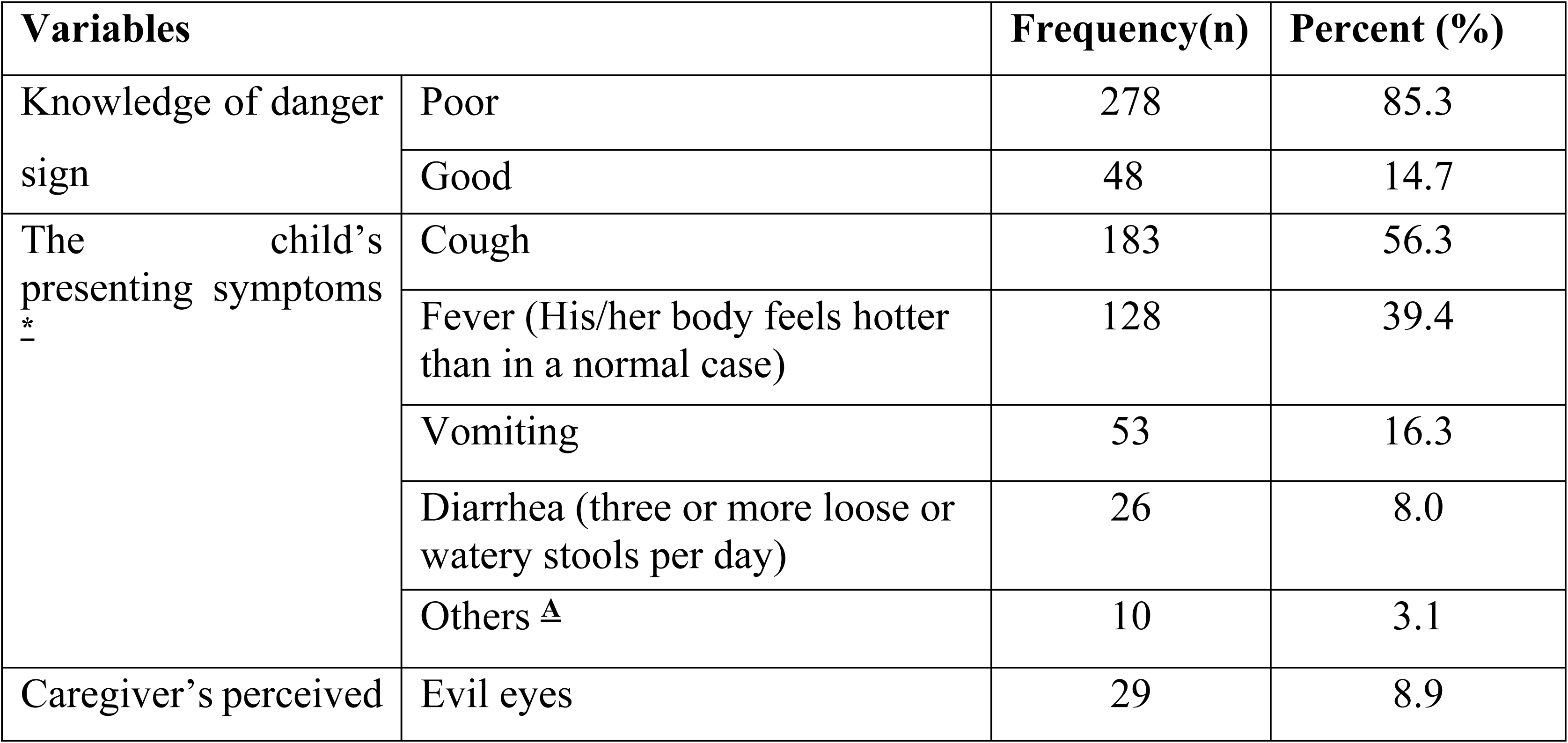
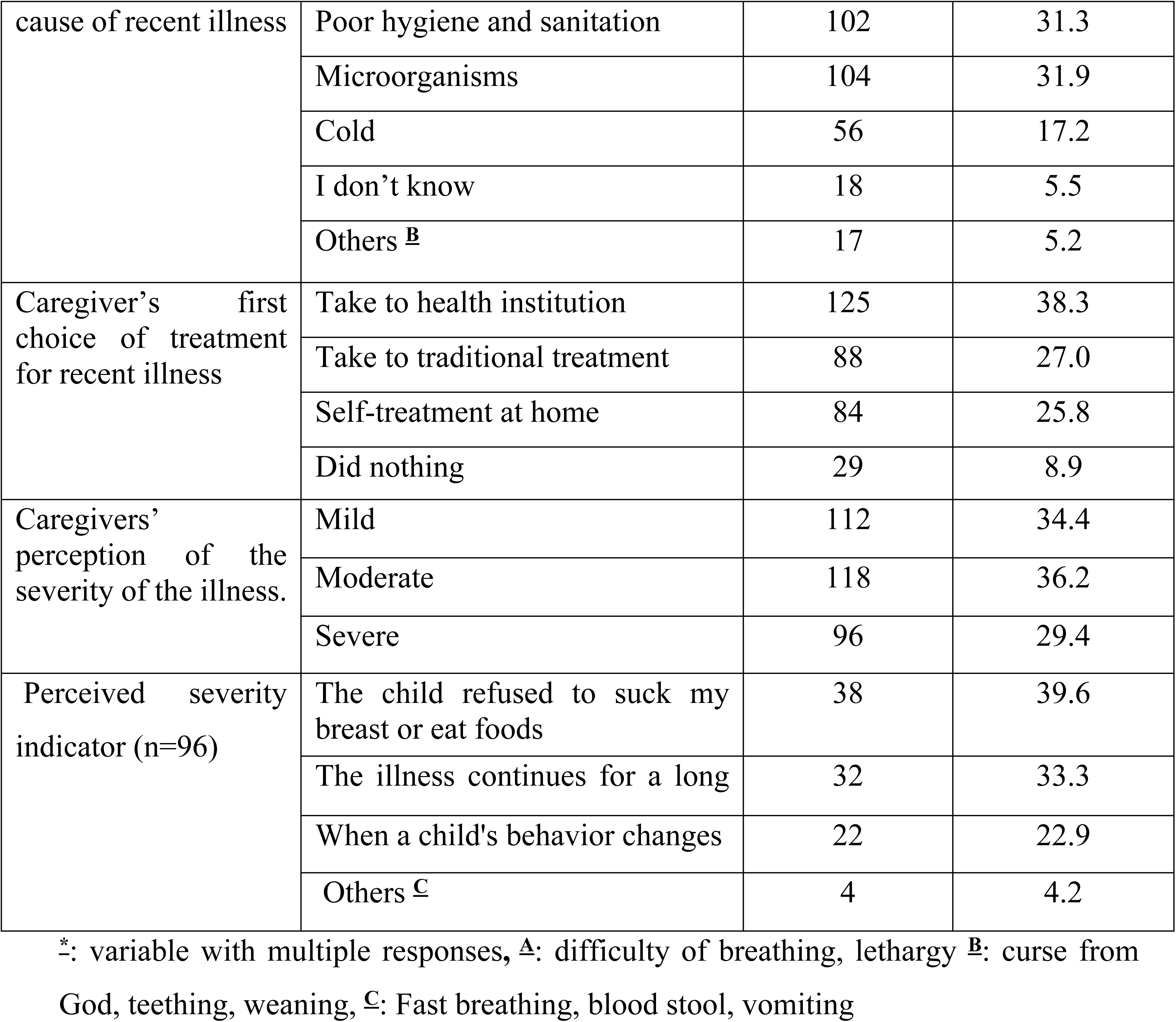
Caregivers’ knowledge and child-illness-related factors in Yem special woreda public health facilities, Southwestern Ethiopia, 2023 (N=326).

### Caregivers’ previous experiences related factors

More than half, 184 (56.4%) participants got information about early treatment seeking and healthcare providers are the main source of information for majority of them. One hundred thirty (39.9%) participants reported that their children encountered illness in the last 6 months. Most of the caregivers, 90 (69.2%) visited a health facility for the illness their child encountered in the last 6 months. ***(Table 3)***

**Table 3:**
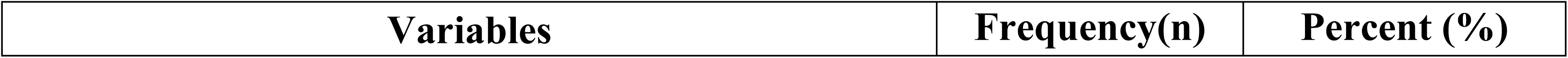
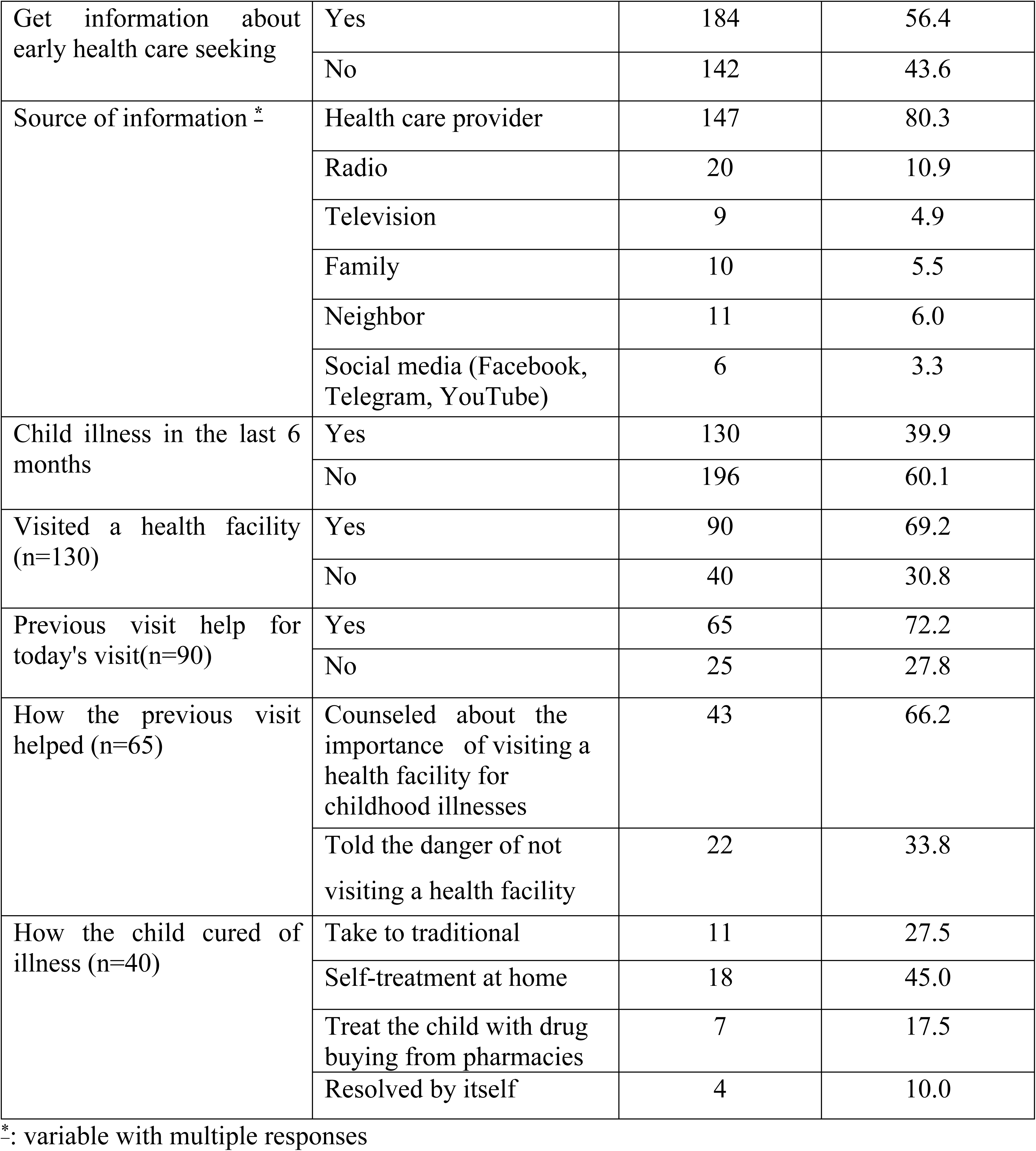
Caregivers’ previous experiences-related factors in Yem special woreda public health facilities, Southwestern Ethiopia, 2023 (N=326).

| Variables | Frequency(n) | Percent (%) |
| --- | --- | --- |

|  |  |  |  |
| --- | --- | --- | --- |
| Get information about early health care seeking | Yes | 184 | 56.4 |
|  | No | 142 | 43.6 |
| Source of information * | Health care provider | 147 | 80.3 |
|  | Radio | 20 | 10.9 |
|  | Television | 9 | 4.9 |
|  | Family | 10 | 5.5 |
|  | Neighbor | 11 | 6.0 |
|  | Social media (Facebook, Telegram, YouTube) | 6 | 3.3 |
| Child illness in the last 6 months | Yes | 130 | 39.9 |
|  | No | 196 | 60.1 |
| Visited a health facility (n=130) | Yes | 90 | 69.2 |
|  | No | 40 | 30.8 |
| Previous visit help for today's visit(n=90) | Yes | 65 | 72.2 |
|  | No | 25 | 27.8 |
| How the previous visit helped (n=65) | Counseled about the importance of visiting a health facility for childhood illnesses | 43 | 66.2 |
|  | Told the danger of not visiting a health facility | 22 | 33.8 |
| How the child cured of illness (n=40) | Take to traditional | 11 | 27.5 |
|  | Self-treatment at home | 18 | 45.0 |
|  | Treat the child with drug buying from pharmacies | 7 | 17.5 |
|  | Resolved by itself | 4 | 10.0 |

\*: variable with multiple responses

### Healthcare access-related factors

In this study, more than half 169 (51.8%) of the participants lived at more than 60 minutes of walking distance from a health facility, in contrast, 157 (48.2%) participants lived within a walking distance of 60 minutes or less from a health facility. The majority, 303 (92.9%) of the participants expressed their preference for seeking healthcare at government health facilities while 23 (7.1%) participants, indicated a preference for private health facilities. Out of 23 participants who prefer private facilities, 18 (78.3%) of them believe treatment in the private is more effective and the remaining being nearby private facilities. Regarding community health insurance, 196 (60.1%) of participants had no community health insurance whereas 130 (39.9%) of participants had community health insurance.

### Decision makers’ behavior-related factors

One hundred seventy-nine (54.9%) of the participants reported that mothers held the primary responsibility for making decisions regarding medical treatment for their children. Most 260 (79.8) of the decision makers do not consume khat and 265 (81.3) do not smoke cigarettes. Nearly half 155 (47.5) of the decision makers reported not consuming alcohol in the last 13 months. ***(Table 4)***

**Table 4:** Decision makers’ behavior-related factors in Yem special woreda public health facilities, Southwestern Ethiopia, 2023 (N=326).

| Variables |  | Frequency(n) | Percent (%) |
| --- | --- | --- | --- |
| Decision maker | Mother | 179 | 54.9 |
|  | Father | 82 | 25.2 |
|  | Both mothers and father | 46 | 14.1 |
|  | Grandparent | 19 | 5.8 |
| Khat chewing | Yes | 66 | 20.2 |
|  | No | 260 | 79.8 |
| Smoking status | Every day | 34 | 10.4 |
|  | Some days | 27 | 8.3 |
|  | Not at all | 265 | 81.3 |
| Alcohol drinking status | Almost every day | 59 | 18.1 |
|  | At least once a week | 82 | 25.2 |
|  | Less than once a week | 30 | 9.2 |
|  | None in the last 13 months | 155 | 47.5 |

### Promptness of healthcare-seeking

About 243 participants reported the signs /symptoms of the illness started more than 24 hrs before the time of seeking care. Therefore, the proportion of delayed healthcare seeking among the study participants was 74.5% (95% CI: 69.8-79.3). More than half 176 (54.0%) of caregivers arrived 2-3 days after symptoms started. The primary reason behind the delay in seeking healthcare was the expectation among 163 (67.1%) participants that the illness would improve by itself. Additionally, other significant reasons for the delay encompassed a reliance on traditional treatments 27 (11.1%), unable to decide by themselves 19 (7.8%), treatment with over-the-counter medication 6 (2.5%), and financial constraints experienced by 28 (11.5%) participants. ***(Figure 1)***

**Figure 1:**
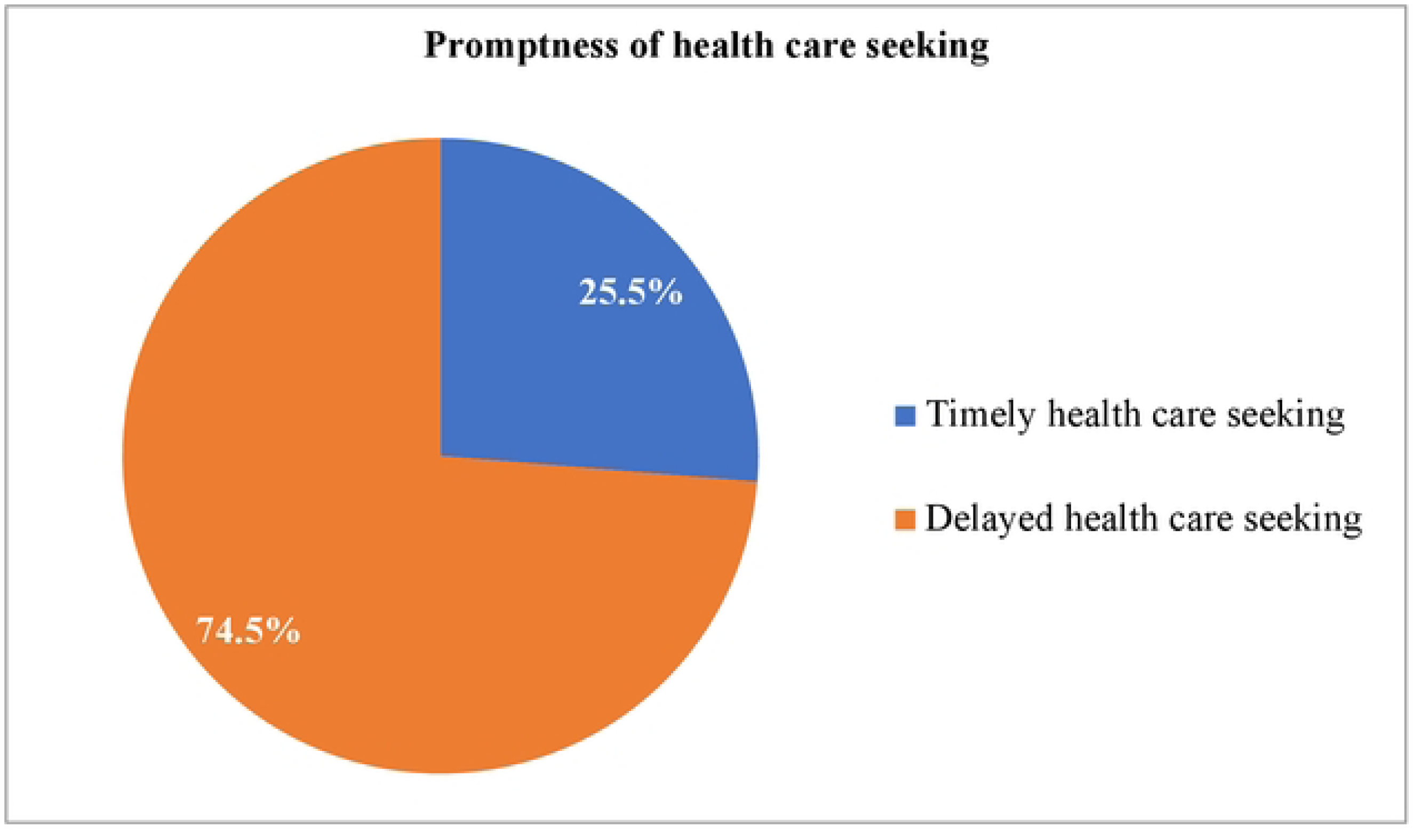
Promptness of healthcare-seeking among caregivers in Yem special woreda public health facilities, Southwestern Ethiopia, 2023 (N=326)

### Factors associated with delay in healthcare seeking for common childhood illnesses among caregivers with under-five children

In this study, the odds of delayed health care seeking were 1.99 times (AOR = 1.99, 95% CI: 1.11-3.57) higher among caregivers who had a child ≥ 12 months than caregivers who had <12 months. The finding also revealed caregivers who reside in rural had 2.41 times (AOR = 2.41, 95% CI: 1.35-4.28) higher odds of delayed health care seeking than those who reside in urban. On the other hand, the odds of delayed care-seeking were 2.98 times (AOR = 2.98, 95% CI: 1.46-6.10) higher for caregivers who used traditional treatment first, and 2.73 times (AOR = 2.73, 95% CI: 1.32-5.63) higher for those who tried self-medication at home first, as compared to going to a health facility first.

In similar ways the finding of this study revealed that caregivers who perceived child illness as mild had 2.64 times (AOR = 2.64, 95% CI: 1.28-5.42) higher odds of delayed health care seeking than caregivers who perceived it as severe. Furthermore, the odds of delayed health care seeking were 1.91 times (AOR = 1.91, 95% CI: 1.07-3.42) higher among care givers who have no community health insurance than those who have community health insurance. ***(Table 5)***

**Table 5:** Bi-variable and multi-variable logistic regression analysis of delay in health care seeking for childhood illnesses among caregivers of under-five children in Yem special woreda public health facilities, Southwestern Ethiopia, 2023 (N=326).

| Variables |  | Delayed healthcare seeking |  | COR (95%CI) | AOR (95% CI) | P-value |
| --- | --- | --- | --- | --- | --- | --- |
|  |  | YES | NO |  |  |  |
| Child age | ≥12months | 157 | 35 | 2.50(1.51-4.16) | **1.99(1.11-3.57)** | **.022\*** |
|  | <12months | 86 | 48 | 1 | 1 |  |
| Caregiver residence | Rural | 187 | 46 | 2.69(1.59-4.54) | **2.41(1.35-4.28)** | **.003\*** |
|  | Urban | 56 | 37 | 1 | 1 |  |
| Family monthly income | Low income (<3000 ETB) | 147 | 43 | 1.84(.69-4.90) | 1.77(.58-5.41) | .317 |
|  | Medium income (3000-75000 ETB) | 83 | 33 | 1.35(.496-3.69) | 1.30(.43-3.96) | .641 |
|  | High income (>7500) | 13 | 7 | 1 | 1 |  |
| Knowledge of danger signs | Poor | 212 | 66 | 1.76(.92-3.38) | 1.47(.70-3.07) | .310 |
|  | Good | 31 | 17 | 1 |  |  |
| First choice of treatment for recent illness | Traditional treatment | 74 | 14 | 3.19(1.62-6.26) | **2.98(1.46-6.10)** | **.003\*** |
|  | Self-treatment at home | 70 | 14 | 3.01(1.53-5.94) | **2.73(1.32-5.63)** | **.007\*** |
|  | Did nothing | 21 | 8 | 1.58(.65-3.86) | 1.01(.38-2.66) | .985 |
|  | Health institution | 78 | 47 | 1 | 1 |  |
| Information about early health care seeking | No | 111 | 31 | 1.41(.85-2.35) | 1.11(.62-1.99) | .715 |
|  | Yes | 132 | 52 | 1 | 1 |  |
| Caregiver's perceived severity of the illness | Mild | 96 | 16 | 3.44(1.76-6.75) | **2.64(1.28-5.42)** | **.008\*** |
|  | Moderate | 86 | 32 | 1.54(.86-2.76) | 1.31(.70-2.46) | .406 |
|  | Sever | 61 | 35 | 1 | 1 |  |
| Distance to the health facility (within walking) | >60 minutes | 131 | 38 | 1.39(.84-2.28) | 1.08(.60-1.94) | .809 |
|  | ≤60 minutes | 112 | 45 | 1 | 1 |  |
| Having community health insurance | No | 159 | 37 | 2.35(1.42-3.91) | **1.91(1.07-3.42)** | **.028\*** |
|  | Yes | 84 | 46 | 1 | 1 |  |

1: Reference category, AOR: Adjusted odd ratio, COR: Crude odd ratio, CI: confidence interval, Bold\*-P value <0.05.

## DISCUSSION

In this cross-sectional study, it was detected that 74.5% (95% CI: 69.8-79.3) of caregivers delayed seeking healthcare for common childhood illnesses in children under the age of five. These findings indicate a substantial proportion of caregivers exhibit a delay in seeking necessary healthcare.

The current finding is consistent with previous studies conducted in the Aneded district of Northern Ethiopia, and Addis Ababa where the proportion of delayed healthcare seeking for common childhood illnesses was reported at 73% and 73.3%, respectively (<u>32</u>, <u>33</u>). This consistency in findings could be attributed to the proximity of caregivers’ perception of the severity of illness within the Ethiopian context. If caregivers perceive the illness as mild, they are more likely to delay seeking care (<u>34</u>).

The results of this study indicate a higher proportion of delayed healthcare-seeking among caregivers compared to the study conducted in Nepal, which reported a proportion of 62.7% (<u>29</u>). This disparity might be due to variations in caregivers’ preferences for healthcare facilities as evidence shows caregivers utilizing public health facilities are more likely to delay seeking medical care compared to those utilizing private health facilities (<u>36</u>). The findings of the present study indicated a significant preference among caregivers for public health facilities compared to the previous study.

Similarly the current study is higher than the study conducted in Bhubaneswar, where the proportion was 15.77% (<u>30</u>). This disparity could potentially be attributed to differences in awareness creation efforts. In the present study area, it was observed that the local institution did not conduct regular organization of role plays and awareness camps, an observation which differs from the practices noted in the previous study area (<u>30</u>).

In contrast, the present study revealed a lower proportion of delayed healthcare seeking when compared to previous studies conducted in the Jeldu district, Oromia region, where a proportion of 86.3% was reported (<u>31</u>). The variation observed may be attributed to the difference in educational status among caregivers, as evidence shows more delayed healthcare-seeking among caregivers with lower educational levels (<u>19</u>). The earlier study documented a notably greater percentage of caregivers with primary education or no formal education in comparison to the present study (<u>31</u>).

The proportion of delayed healthcare seeking in current study is also lower than a study conducted in Cameroon, which reported a proportion of 88.1% (<u>28</u>). This disparity could be attributed to differences in the timing of the studies. Previous research was carried out during the COVID-19 (Coronavirus Disease 2019) pandemic, which has resulted in widespread disruptions to healthcare systems. These disruptions include decreased access to healthcare facilities, concerns about contracting the virus, diminished financial capacity to afford healthcare, and alterations in healthcare-seeking behaviors. (<u>44</u>, <u>45</u>).

The findings of this study revealed caregivers of children ≥ 12 months were more likely to delay seeking healthcare compared to caregivers of infants below 12 months. This observation aligns with studies conducted in Nekemte (<u>34</u>) and Tanzania (<u>46</u>). The underlying reasons for this may be attributed to caregivers often prioritize the care of younger infants, holding the belief that children older than one year of age are more resilient to illness (<u>34</u>). Furthermore, caregivers may possess an awareness that younger children are more susceptible to severe illnesses, which could contribute to their prompt healthcare-seeking behavior for infants below 12 months (<u>47</u>, <u>48</u>).

The present study revealed a higher likelihood of healthcare-seeking delay among caregivers residing in rural areas compared to those in urban settings, which aligns with findings from previous studies conducted in Nekemte town (<u>34</u>), Bahir Dar city (<u>35</u>), Hawassa (<u>49</u>), Kaduna Nigeria (<u>50</u>), and India (<u>51</u>). This might be due to variations in the accessibility of public health services, resulting from geographical distance and media exposure (<u>52</u>). Moreover, it could also be due to challenges posed by poverty and limited healthcare access in rural areas (<u>53</u>). In contrast, urban households tend to benefit from better accessibility to healthcare services (<u>54</u>).

On the other hand, the findings of this study showed that caregivers who lack community health insurance are more likely to delay seeking healthcare in comparison to those who have community health insurance. This result is in line with previous studies conducted in Nekemte and Debre Markos town (<u>34</u>, <u>55</u>). The reason might be being uninsured creates concerns about the financial burden associated with seeking healthcare as community-based health insurance significantly reduces healthcare costs (<u>34</u>, <u>56</u>).

The findings of this study showed that the odds of delayed healthcare-seeking are increased when caregivers resort to traditional medicine or self-medication. This result aligns with previous studies conducted in Bahir Dar and Nekemte (<u>34</u>, <u>35</u>). The underlying reasons for this can be attributed to cultural beliefs that promote the use of home remedies and traditional medicine as the initial and acceptable form of treatment, as well as the community’s adherence to religious beliefs and practices (<u>36</u>). Moreover, caregivers’ perceptions regarding the effectiveness of traditional treatments may contribute to this trend (<u>57</u>). Home remedies are often perceived as cost-effective and easily obtainable (<u>58</u>).

Consistent with a study conducted in Nekemte (<u>34</u>), the present study identified that caregivers who perceived their child’s illness as mild exhibited a higher likelihood of delayed healthcare-seeking, compared to those who perceived the illness as severe. This might be due to caretakers tend to seek care from health facilities more frequently when they believe their children are seriously ill, as they want to prevent any potential complications (<u>29</u>). Moreover, this phenomenon may be ascribed to caregivers’ conviction that their child’s illness will spontaneously ameliorate over time, prompting them to adopt a “wait-and-see” approach until further symptoms manifest (<u>16</u>).

## Data Availability

All relevant data are within the manuscript and its Supporting Information files.

## Declarations

### Ethics approval and consent to participate

Provided in Methods and Material section of the manuscript.

### Consent for Publication

Not applicable

### Availability of Data and Materials

The authors presented data in the main paper and will be available upon request from the corresponding authors.

### Authors’ contribution

KF - conceived research idea, questionnaire design, conducted the fieldwork, analyzed the data, interpretation of results, results and discussion writing.

GAH - contributed to the revision of the research and questionnaire design, supervision of fieldworks, involved in statistical analysis, and development of the manuscript.

TD - contributed to the revision of the research and questionnaire design, supervision of fieldworks, involved in statistical analysis, and development of the manuscript.

All authors read and approved the final manuscript.

## Acknowledgment

The authors are grateful to Jimma University, Institute of Health, Faculty of Health Sciences, School of Nursing. We also want to express our sincere appreciation to the Yem special woreda health office and the stakeholders in health facilities for their cooperation. We would like to thank the data collectors and supervisors. Finally, we are thankful to the study participants who generously volunteered their time.

